# Stochastic Compartmental Modelling of SARS-CoV-2 with Approximate Bayesian Computation

**DOI:** 10.1101/2020.03.29.20046862

**Authors:** Vedant Chandra

## Abstract

In this proof-of-concept study, we model the spread of SARS-CoV-2 in various environments with a stochastic susceptible-infectious-recovered (SIR) compartmental model. We fit this model to the latest epidemic data with an approximate Bayesian computation (ABC) technique. Within this SIR-ABC framework, we extrapolate long-term infection curves for several regions and evaluate their steepness. We propose several applications and extensions of the SIR-ABC technique.

## 1. EPIDEMIC MODEL

The SIR model (Kermack & McKendrick 1927) traces 3 trajectories in phase space: susceptible (S), infectious (I), and recovered members of the population (R). The transmission rate *β* represents the number of disease transmissions per unit time, per infected host. The recovery rate *γ* is simply the number of recoveries per unit time. The disease lifetime is exponential, with a wait time scaling as *e*^*−γt*^. The expectation of disease dura-tion is hence 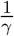. These parameters govern the disease model with the following differential equations:

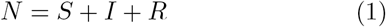

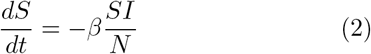

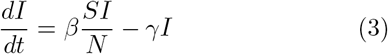

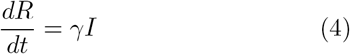

We use an implementation of the Gillespie algorithm (Gillespie 1977) to generate stochastic trajectories of *S, I*, and *R* from these differential equations.

## 2. APPROXIMATE BAYESIAN COMPUTATION

Armed with the ability to generate stochastic infection and recovery curves from starting parameters, we turn to fitting the starting parameters from real-world epidemic data. Since the models are stochastic in nature, there isn’t a simple analytical form that we can minimize. Additionally, rather than fitting for only the parameters themselves, we would also like to quantify how certain we are about those parameters. We therefore employ an approximate Bayesian computation (ABC) technique to compare our simulations to observations and recover the posterior distributions of *β* and *γ* (Figure 1). This technique was previously used to fit initial mass functions to nearby galaxies (Gennaro et al. 2018).

**Figure 1.**
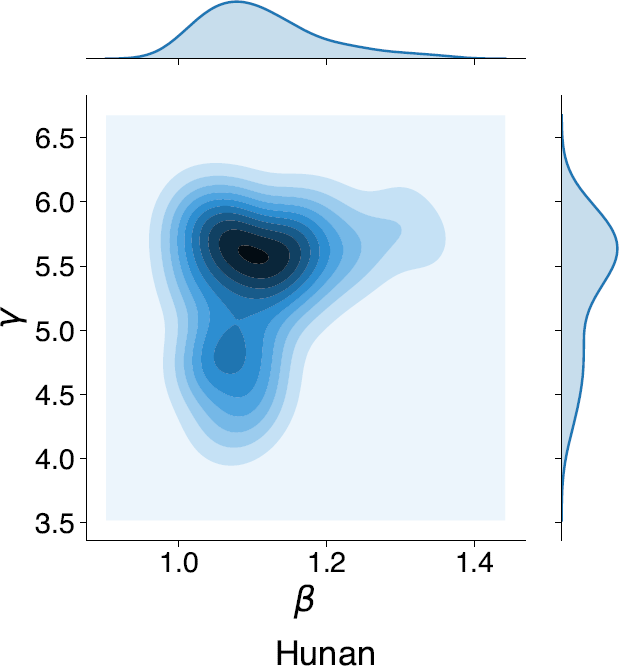
An advantage of the SIR-ABC method is the ability to fully capture the covariate joint distribution between fitted model parameters.

The general goal of ABC is to sample the posterior distributions of simulation parameters such that the simulations match the observed data. In practice, it is impossible for simulations to exactly match data due to noise and ill-posed models. Additionally, if the observable space is continuous, then the probability of simulations exactly matching observations is exactly zero. Therefore, we define some distance *d* between simulations and observations, as well as a tolerance *ϵ*. We accept those parameters who produce simulations are *d < ϵ* away from the observed data. By initially sampling from the prior distributions of the parameters and iteratively shrinking the tolerance *ϵ* up to some stopping criterion, we ‘shrink’ the prior into the posterior.

The Dong et al. (2020) epidemic data consists of a 2-dimensional time series comprising of the number of confirmed cases and the number of recovered cases per day (*R*). We subtract these two quantities to derive the number of infectious cases per day, *I*. Given a simulated epidemic and the observed data, we quantify the difference between both the infectious and recovered population curves to obtain a distance

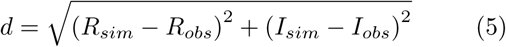

Rather than *a-priori* assuming the initial susceptible population *S*, we marginalize over it as a nuisance parameter in our ABC procedure. Therefore, our ABC algorithm fits for three parameters: *β, γ*, and *S*.

We use the pyabc package in Python (Klinger et al. 2018) for our ABC procedure. We employ a simple particle filter algorithm (sequential Monte Carlo) that accepts or rejects sampled particles based on the selection criterion *d < E*, until *p* particles have been accumulated. The first iteration samples uniform priors on each parameter, and each subsequent iteration samples the posterior of the previous iteration. We shrink *ϵ* by setting *ϵ*_*i*_ of the *i*^*th*^ iteration equal to the median of all the sampled distances *d* from the (*i −* 1)^*th*^ iteration.

As the parameters converge to their posterior, the shrinkage of *ϵ* slows down, and the sampler has to reject progressively more particles in order to accumulate *p* particles with *d < E*. We choose a stopping criterion such that the acceptance ratio (number of total particles sampled in order to accumulate *p* valid particles) is 1%. We find that the models are well-converged at this point, and sampling further does not improve the parameter posteriors. The root mean square difference defined in Eqn. 2 is around *d ∼* 10 for the converged models. Each fit takes *∼* 25 minutes to complete on a regular laptop computer.

## 3. RESULTS

We fit our model to the 12 provinces in China worst affected by SARS-CoV-2, with the exception of Hubei due to the lack of early-stage data there. We recover posterior densities of *β, γ*, and the number of susceptible citizens *S* (Figure 2). We present epidemic curves with our model simulations overlaid for all regions in the appendix.

**Figure 2.**
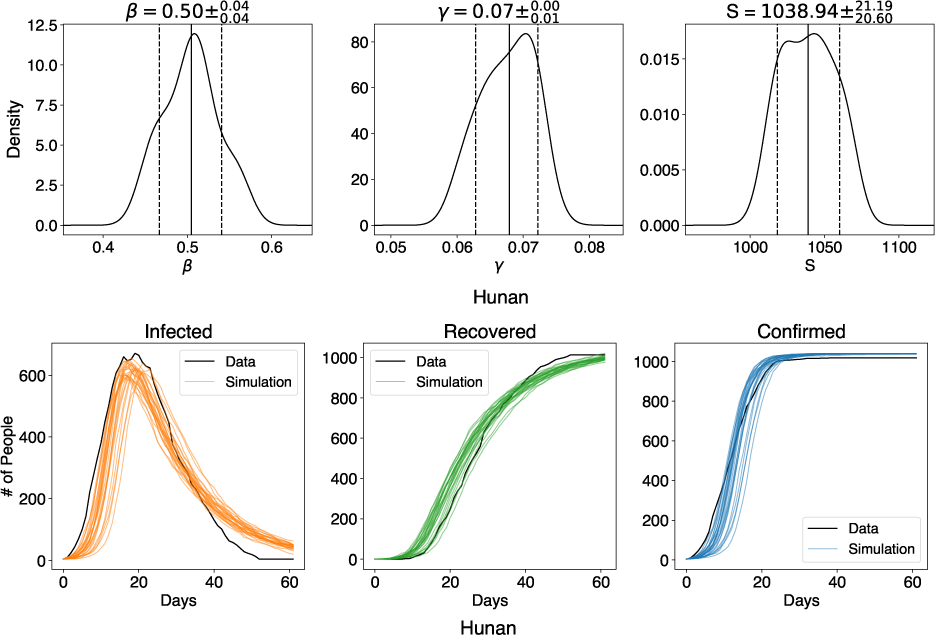
Above: Posterior distributions of our fitted epidemic parameters for the Hunan province. Below: Observed epidemic curves, along with 25 realizations of the best-fit model.

For most provinces, there is an excellent agreement between the SIR-ABC model and the total number of confirmed cases. The fit is less perfect for the individual infected-recovered curves. This is to be expected, since the real-world obviously does not truly follow an SIR model. There are various externalities like spatial effects and government/healthcare responses. Our simple SIR model also lacks vital statistics like births and deaths. For a fatal illness like SARS-CoV-2, it would be valuable to add these parameters to the model. However, for the purpose of this proof-of-concept study, we estimate that adding these parameters will negligibly affect the goodness-of-fit of the total confirmed cases (Chen & Li 2020).

We extrapolate the model for each region by allowing it to run until no active infections remain (Fig. 3). We find a consistent extrapolated infection profile for all the provinces under study. This indicates a similar level of government response after the first infections were reported, despite differing population sizes in each region. We quantify the ‘steepness’ of the infection curve by dividing the maximum number of active infected patients by the total length of the extrapolated infection curve, i.e. the duration of the epidemic. We compare the steepness of different Chinese provinces in Fig. 4.

**Figure 3.**
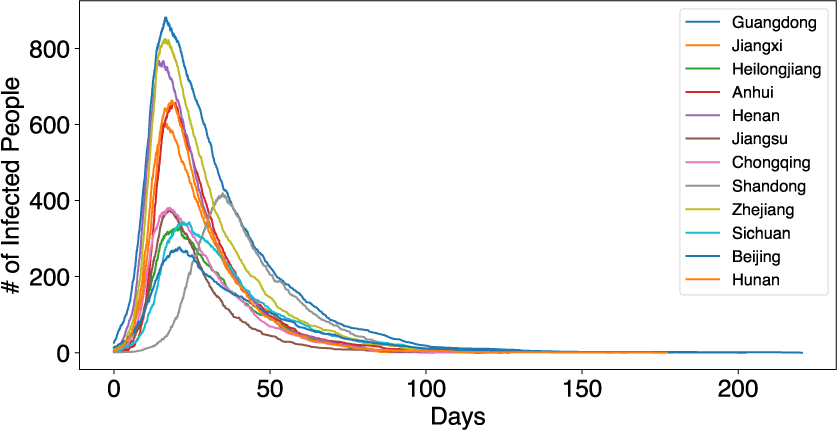
Extrapolated infection curves for the 10 worst-affected Chinese provinces. We allow the epidemic solution to continue until no active infections remain.

**Figure 4.**
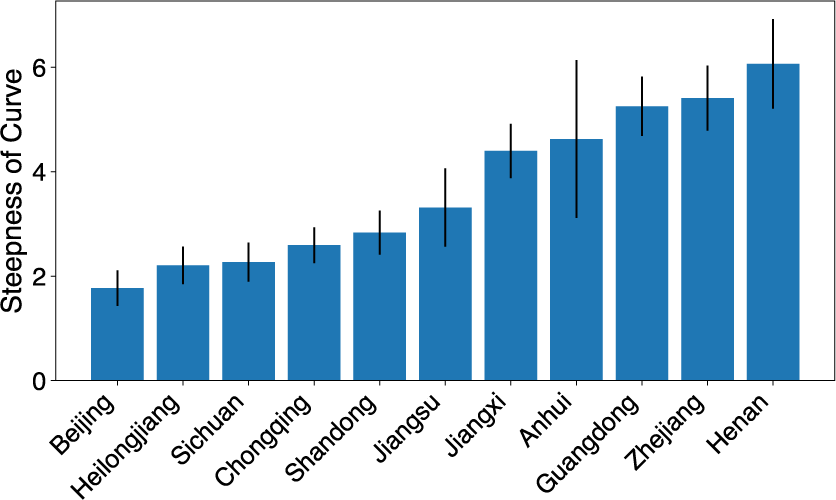
Relative ‘steepness’ of the extrapolated infection curves in Fig. 3.

We find a strong correlation (*p <* 0.01) between the steepness of the infection curve and the fitted initial number of susceptible patients. This is likely not a significant finding, but rather an intrinsic collinearity between these measures.

## 4. DISCUSSION

In this proof-of-concept study, we apply approximate Bayesian computation to fit stochastic epidemic models to real world data. We encourage researches to improve and adapt these methods to other problems.

An interesting extension of our analysis would be characterizing the reproduction rate *R*_0_ of different regions. However, we use a non-standard parameterization of the SIR model for the benefit of our ABC optimization. Therefore, our derived *R*_0_ = *β/γ* lacks interpretability and cannot be compared to other studies. We invite other researchers to repeat our analysis with the standard SIR parameterization.

Additionally, whilst parameter fits are poorly constrained in populations where the infection has not already peaked, it would be interesting to explore epidemic forecasting on those datasets. The Gillespie algorithm can be optimized to work faster with larger numbers of patients. Our parameterization of the SIR model can also be modified to include vital statistics like births and deaths. ABC generalizes well to these higher-dimensional parameter spaces. Specific to SARS-CoV-2, age-structured models would also be a valuable development, as would models that include vaccinations and acquired immunity.

## Data Availability

This study uses the data repository for the 2019 Novel Coronavirus Visual Dashboard operated by the Johns Hopkins University Center for Systems Science and Engineering (JHU CSSE), supported by ESRI Living Atlas Team and the Johns Hopkins University Applied Physics Lab (JHU APL)

https://github.com/CSSEGISandData/COVID-19

## APPENDIX

**Figure 5.**
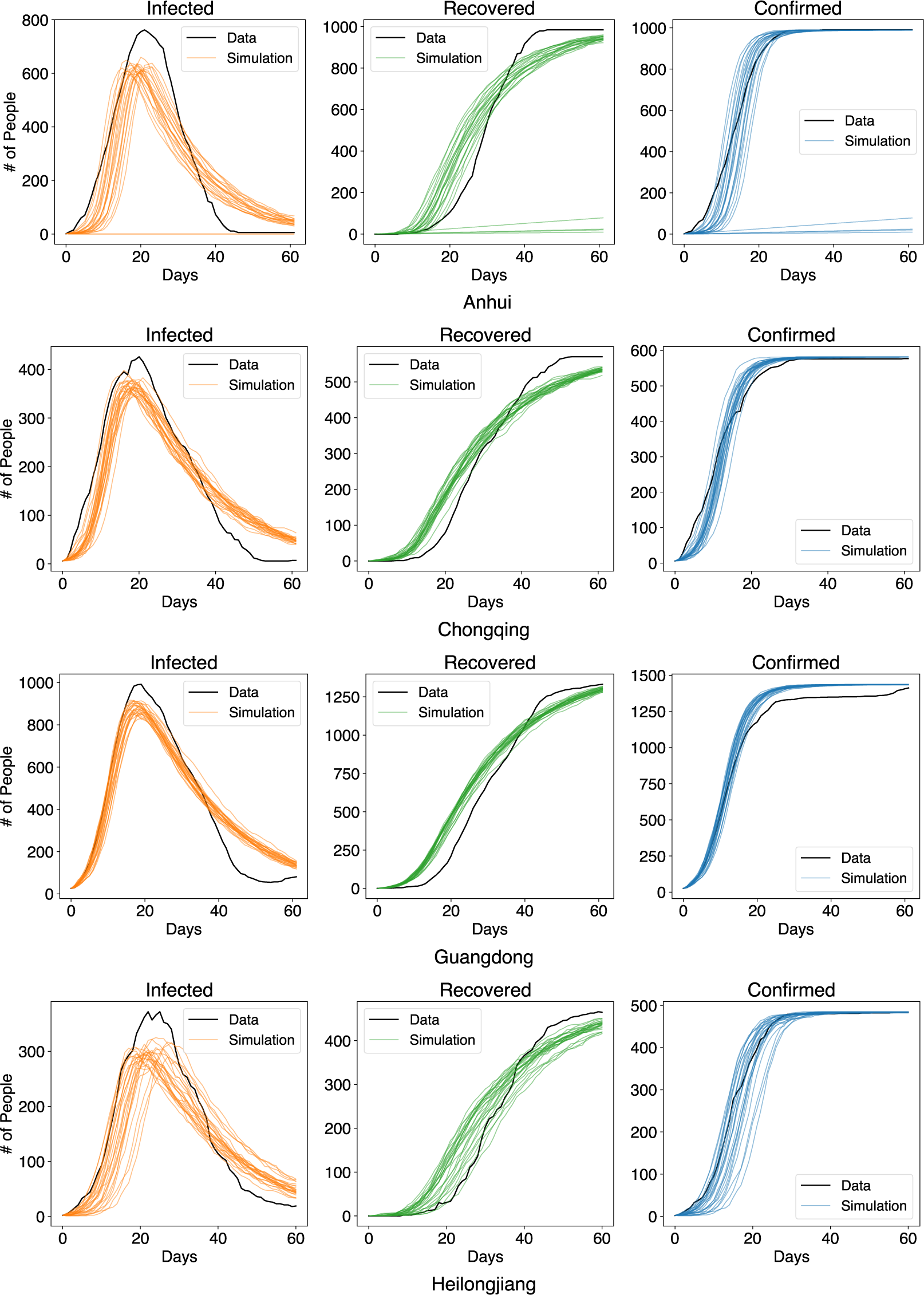
Observed epidemic curves, along with 25 realizations of our best-fit SIR-ABC model for 4 Chinese provinces.

**Figure 6.**
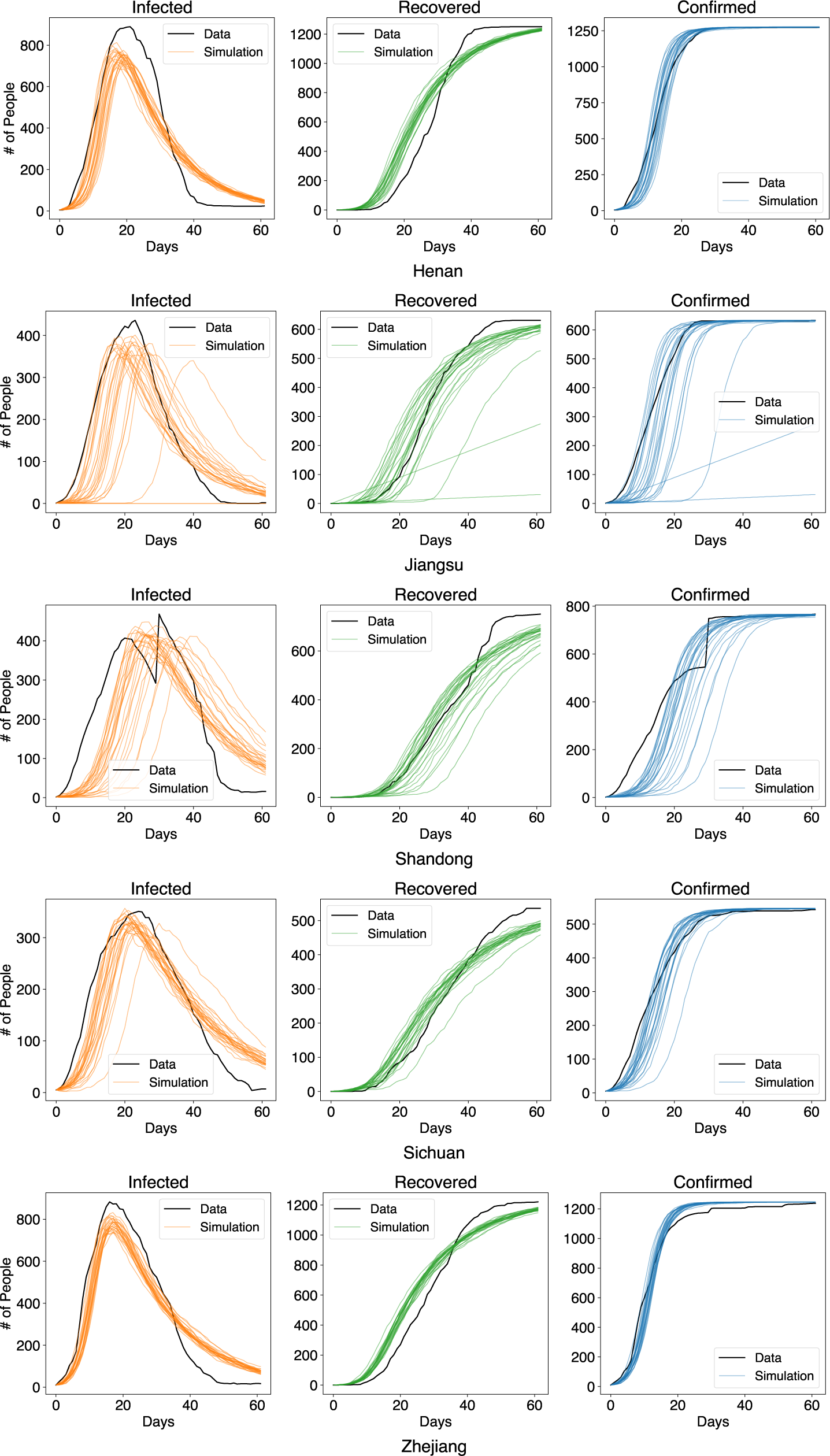
Observed epidemic curves, along with 25 realizations of our best-fit SIR-ABC model for 5 more Chinese provinces.

